# Timeliness in canine rabies outbreak report and response

**DOI:** 10.1101/2025.09.30.25336678

**Authors:** Elvis W. Díaz, Alejandra Dávila-Barclay, Antuannete Vela, Laura D. Tamayo, Valerie A. Paz-Soldan, Claudia Mena, Ricardo Castillo-Neyra

## Abstract

**Introduction:** Timely response to zoonotic disease outbreaks is crucial to minimizing their impact on the health and wellbeing of people and animals. Monitoring and improving the speed of response, using established outbreak milestones and timeliness metrics, is key to preventing further spread. Despite training and a decade of expertise in case response, persistent delays in these processes continue to hinder effective outbreak responses. Dog- mediated rabies, a classic One Health challenge, remains a significant public health concern in many parts of the world. In Arequipa, Peru, rabies has re-emerged and caused recurrent outbreaks over the past decade, despite substantial control efforts. Addressing the drivers of these delays is vital for improving rabies control. In this study, we use dog rabies as a model to examine delays in zoonotic outbreak response, as well as potential solutions, applying and adapting the Outbreak Milestones and Timeliness Metrics framework. We calculated the timeliness of rabies surveillance and response activities in Arequipa from 2015 to 2024 and identified key barriers contributing to delays.

**Methods:** Using canine rabies outbreak response reports from 2015 to 2024 in Arequipa, Peru, we classified One Health outbreak milestones pertaining to the report and response system and calculated timeliness metrics between key processes. Kruskal-Wallis and Dunn tests were applied to measure delays by process, year, and micro-health network. Qualitative analysis of narrative field reports identified underlying causes of delays.

**Results:** The longest delay occurred between the detection of behavior change in dogs and initial reporting, averaging 2.52 days (SD = 2.28), while the shortest delay was between diagnosis and outbreak control, averaging 0.76 days (SD = 0.92). Intermediate delays were observed between report and sample shipment and diagnosis and control with averages of 1.28 days (SD = 1.30) and 1.09 days (SD = 1.38), respectively. Delay-related barriers included dog owners’ lack of awareness and reluctance to report symptoms, logistical challenges in sample collection, and delays in implementing outbreak control measures. Additional barriers included confusion about reporting procedures and reluctance from dog owners to share information about their dogs which hindered timely response.

**Conclusions:** Delays in rabies reporting and response in Arequipa are primarily driven by slow reporting following behavior changes in dogs and logistical problems with sample collection and control and containment measures. Although diagnosis and outbreak responses are comparatively more efficient, inconsistencies remain. Strengthening community awareness and engagement for reporting, improving intersectoral coordination between health authorities, and optimizing logistics are critical to reducing delays and improving outbreak control.

## Introduction

Timely implementation of public health responses to zoonotic disease outbreaks is critical to mitigate their impact on the health and well-being of humans, animals and environmental health, as well as to minimize broader economic consequences affecting livelihoods. Delays in emergency response have long been studied; notably, the Three Delays Model of Maternal Mortality identified critical decision points where delays can worsen outcomes, in that specific case, on maternal deaths. In recent years, with the increasing risk of infectious diseases outbreaks escalating into epidemics or pandemics, the need for coordinated and timely responses have increased. This has led to the development of timeliness-based frameworks to monitor and evaluate the effectiveness of surveillance and early-response activities. Recognizing the complexity of zoonotic disease threats and our global interconnectedness, such metrics have been adapted to encompass all components of the One Health triad, resulting in initiatives like the Salzburg Global Seminar’s Statement on Metrics for One Health Surveillance which was convened by a quorum of experts in global outbreak response. These frameworks propose measuring time between 11 milestones or key processes of an outbreak response: Prediction, Prevention, Outbreak Start, Detection, Notification, Verification, Diagnostics, Response, Communication, Outbreak End, and After-Action Review. Reducing time intervals between milestones is critical to obtaining optimal public health outcomes. Evaluating real-world applications of this framework is essential to identify sources of delays in various zoonotic disease responses. Rabies, a classic example of a One Health challenge, offers a valuable model to study these metrics in action.

Dog-mediated rabies continues to be a neglected zoonotic disease (NZD) and a major global health problem, with a fatality rate approaching 100% and an estimated 59,000 human deaths annually. In Peru, canine rabies maintained endemic foci in various regions throughout the 20th century until the implementation of the Action Plan for the Elimination of Urban Rabies, which successfully achieved control by the early 2000s. In 2013, Arequipa, one of the largest cities in the Peruvian Andes, was part of the 88% of Peruvian territory declared free of canine rabies. The neighboring Peruvian department of Puno, which shares a border with Bolivia, continued to report cases. In 2015, a rabies-positive dog was detected in Arequipa, marking the first documented reintroduction of canine rabies in Peru and Latin America. Since then, rabies-positive cases have been reported consistently, averaging around one case per week.

In response to the reintroduction of canine rabies and with the goal of elimination in Arequipa, the Ministry of Health (MOH) has implemented different strategies, primarily focusing on mass canine rabies vaccination campaigns (MDVC, known as VanCan), alongside surveillance and control measures. A key component of the response is targeted outbreak control, involving the epidemiological investigation following a confirmed canine rabies case. The primary objective of this approach is to prevent secondary cases in animals and reduce the risk of virus transmission to humans through contact tracing for both humans and dogs following a positive case confirmation, as well as ring vaccination. These combined strategies represent the core efforts to control and contain the disease within the city.

Despite ongoing efforts in containment of positive cases and community-based health promotion to enhance case reporting, delays in the reporting of rabid dogs within communities in Arequipa persist. Despite the commitment of local health officials to strengthen the canine rabies surveillance program, response delays have been observed at various stages of the control response. These delays may increase morbidity and mortality, hindering public health efforts. Contributing factors include community-level and public health system-level barriers – ranging from human and logistical to economic factors– that hinder timely reporting, sample processing, implementation of control measures, and care for at-risk individuals.

This study aimed to identify outbreak milestones within the rabies surveillance and response activities conducted in Arequipa between 2015 and 2024, and to measure and monitor the timeliness metrics of the associated processes. We also analyzed narrative field reports of response actions for all cases to examine the underlying causes and barriers contributing to delays in surveillance and response system. By applying the Outbreak Milestones and Timeliness Metrics framework, we also assessed potential solutions to improve response efficiency. This study serves as a practical model of the framework in a real-life zoonotic outbreak, with the goal of improving strategies to reduce delays and improve outbreak management.

## METHODS

### Ethics statement

The study protocol (Project number 210370) was approved by the ethical review committee at Universidad Peruana Cayetano Heredia (Approval number: 314-29-23).

### Study site

This study included rabies cases from all 17 districts of the city of Arequipa and one district (Majes) of the Caylloma province of Arequipa state, Peru. [Figure 1] Situated at 2,300 meters above sea level, Arequipa had an estimated human population of 1,251,101 in 2024, with approximately 76,204 Majes residents (Gobierno Regional de Arequipa, 2024; INEI). The first canine rabies case in the city was reported in March 2015 (Dirección General de Epidemiología, 2015; Pan American Health Organization (PAHO), 2015, p. 201), and as of December 2024, 392 rabid dogs had been documented. At the time of this study, the Ministry of Health (MOH) estimated a canine population of approximately 256,000 dogs across the study area, with human-to-dog ratios varying by district from 3.19:1 to 6.94:1 (Arequipa Regional Health Management).

**Figure 1.**
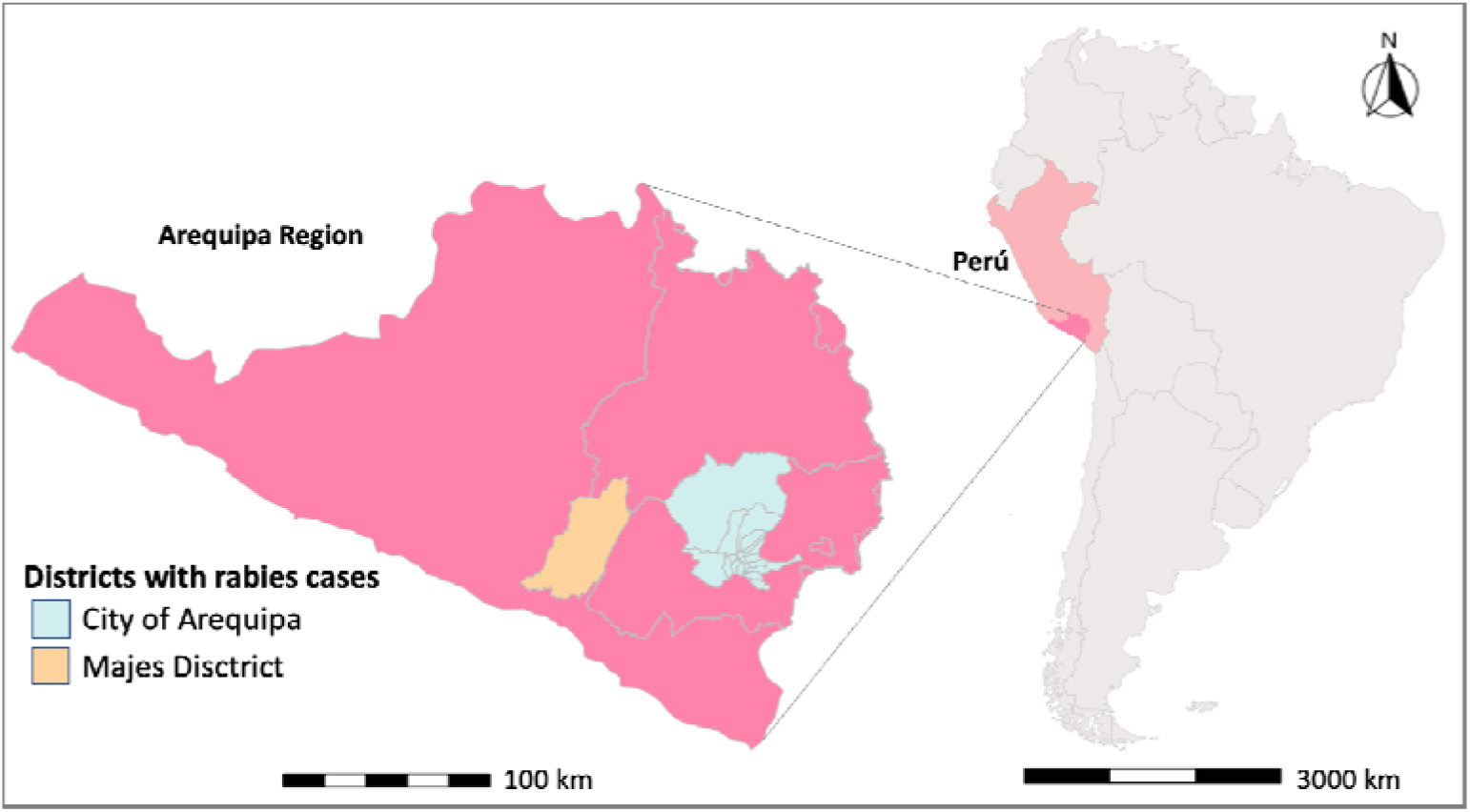
Map of the Arequipa Region, located in southern Peru, showing the location of all districts with dog-mediated rabies cases between 2015 and 2024.

**Figure 2.**
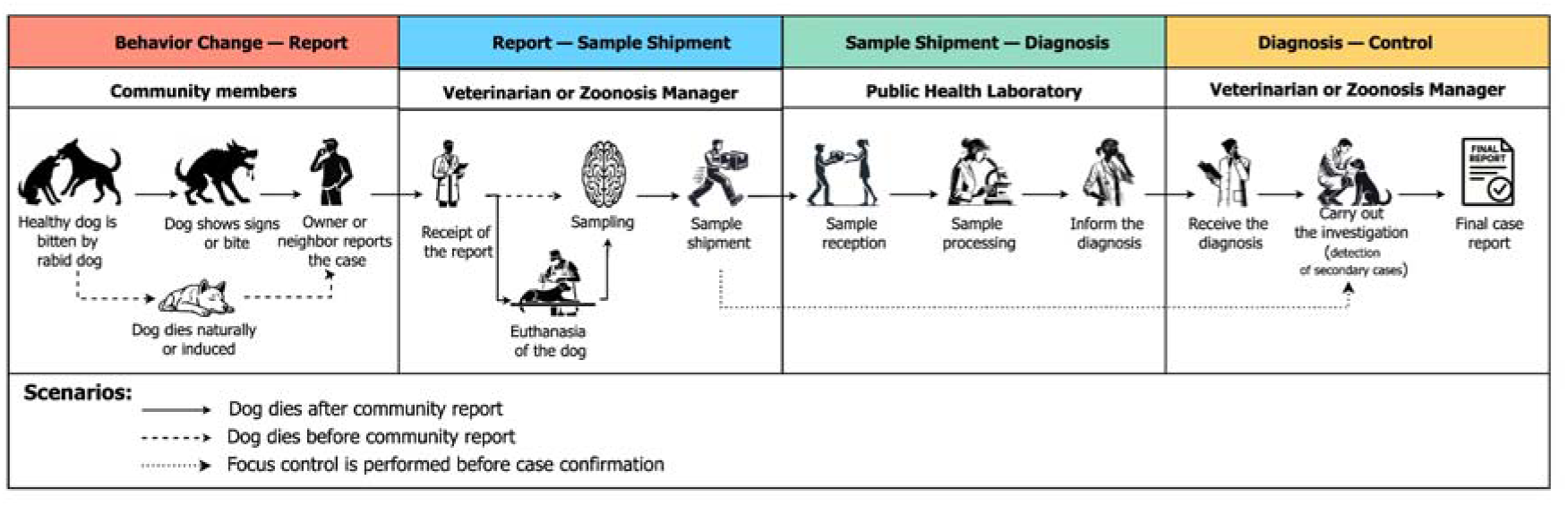
Processes carried out in the canine rabies surveillance and response system in Arequipa. The headings correspond to the processes in the canine rabies outbreak surveillance in Arequipa and the key actors responsible for each step. The solid arrows represent different scenarios in passive and active surveillance and instances in which outbreak investigation is conducted in parallel with laboratory sample processing, before case confirmation by a positive laboratory test result. The images used in this diagram were created using AI tools, specifically DALL·E and ChatGPT.

**Figure 3.**
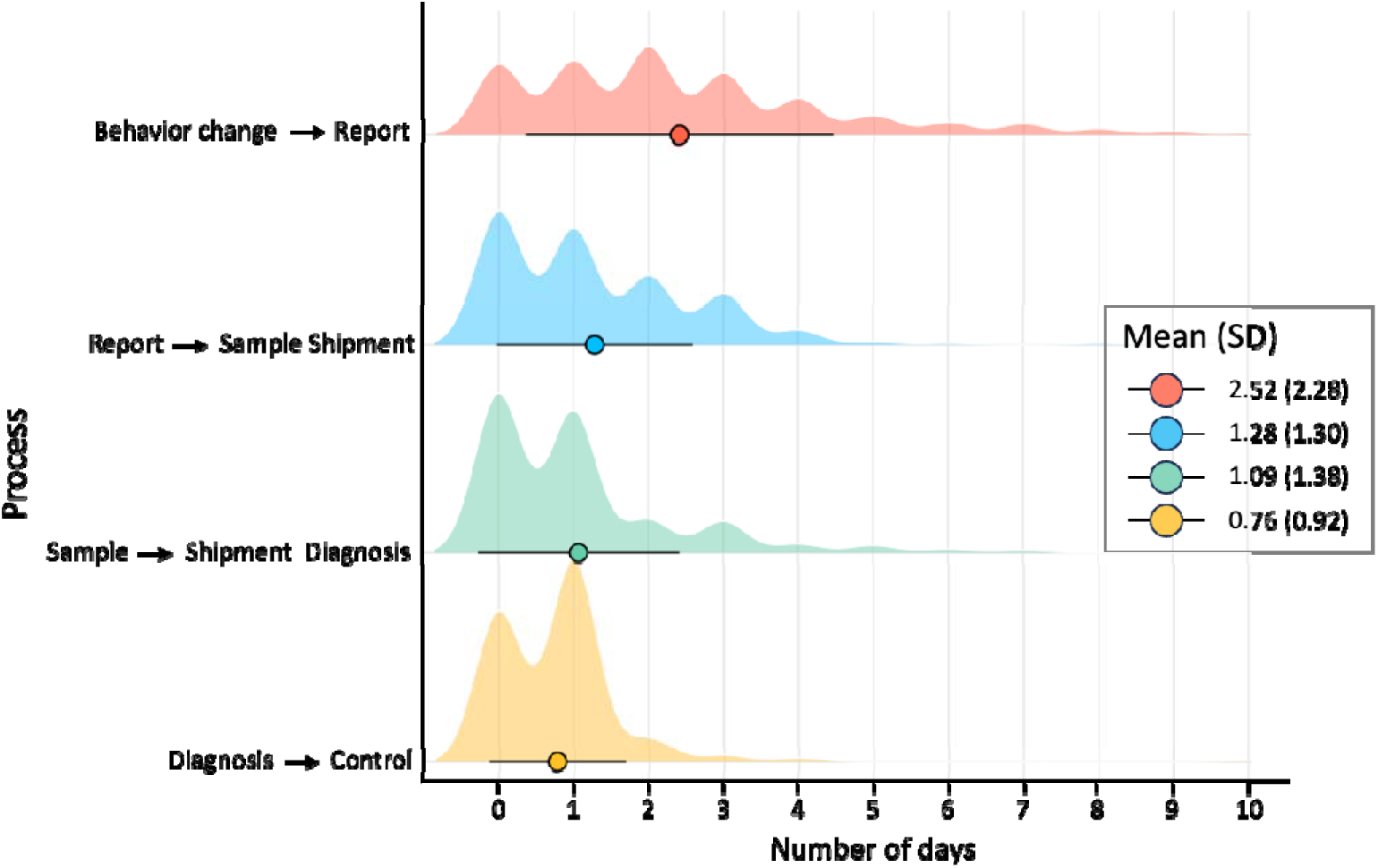
Average days spent between each step in the process of the canine-rabies surveillance system in Arequipa Peru from 2015 to 2024.

Arequipa city has a diverse spectrum of urbanization, migration, and socioeconomic status. The city has a well-defined urban center, with established older neighborhoods and paved sidewalks and roads, as well as access to transport. In contrast, peri-urban areas are rapidly expanding through internal migration and are marked by lower socioeconomic status, higher crime rates, and rugged terrain that hinders mobility and access to services. These areas have fewer community resources and present additional challenges for public health interventions (Castillo-Neyra, Brown, et al.; Buttenheim et al.; Levy et al.).

### Surveillance system

The current National Health Guideline for the Prevention, Surveillance and Control of Human Rabies in Peru outlines two types of rabies surveillance: passive and active. Passive surveillance relies on community-initiated reporting of events to health authorities, such as behavioral changes, biting incidents, or unexplained canine deaths, without proactive intervention from the health system. The passive surveillance system heavily depends on community vigilance and cooperation with health authorities. In contrast, active surveillance involves targeted field investigations by health system personnel, such as the systematic search and collection of brain samples from deceased dogs found in areas commonly used by animals for movement and feeding, such as dry streambeds (inactive water channels) called “torrenteras”.

### Targeted outbreak response activities

A key component of canine rabies control is targeted outbreak response, sometimes referred to as focal control procedures, focus control, or “control de foco” in Spanish. The activities involve conducting epidemiological investigations of confirmed rabies cases in dogs and implementing ring vaccination within a 500-meter radius of the index canine case. The main goal of focal control procedures is to prevent secondary cases in both animals and humans. Focal control typically also includes other activities, such as community education and the systematic identification and management of individuals potentially exposed to the rabid animal.

### Data collection

The study analyzed data from 319 confirmed canine rabies case reports between 2015 and 2024 in Arequipa’s 17 districts and one district in Caylloma province. Case reports were collected from two primary sources. The first source includes epidemiological investigations conducted by the MOH between 2015 and 2018 of suspected canine rabies cases, following its reintroduction in Arequipa. The second source comprises case reports from 2019 to 2024 generated by our research team at the Zoonotic Diseases Research Laboratory (ZDRL) of the One Health Research Unit at Universidad Peruana Cayetano Heredia (UPCH), in close collaboration with the Arequipa Regional Health Directorate (GERESA) of the MOH. During this period, the ZDRL participated in the surveillance and outbreak response to positive cases of canine rabies. All (n=319) cases were laboratory confirmed using direct immunofluorescence (DIF), PCR, or the mouse inoculation test.

Our research team’s reports are primarily narrative field-based accounts, often written in diary or logbook style, detailing the circumstances of each case as observed during targeted outbreak control activities. These also included information from interviews conducted as part of the outbreak investigations. Interviewees ranged from individuals who initially reported the suspected rabid dog, to health inspectors (health center veterinarian or zoonosis manager) who responded to the event notification. These unstructured key informant interviews varied in number and length, aimed to gather as much detailed and contextual information about each case to reconstruct its origins and dynamics. The ZDRL reports (2019-2024) include more detailed narrative descriptions of the events observed and reported compared to those by MOH, however, both MOH (2015-2018) and ZDRL (2019-2024) consistently documented the same set of variables using standardized definitions. These variables included: date of symptom onset; date of initial reporting of reported behavior change or start of symptoms by the owner or community member; and dates corresponding to key response milestones, such as date of case verification, sample shipment, laboratory confirmation, and initiation of control activities. Incomplete reports and those not submitted by the health centers to the Arequipa Health Network (Red de Salud) during 2015, 2016, and 2017 were excluded from the analysis.

### Classification of the Outbreak Surveillance and Response Process within the Outbreak Milestones Framework

We classified the outbreak response process using the Outbreak Milestones Framework, as defined in the Salzburg Statement on Metrics for One Health Surveillance (Salzburg Global Seminar, 2020) (Table 1). Each of the 11 milestones marks a key stage in the outbreak timeline, such as symptom onset, community reporting, sample collection, laboratory confirmation, and initiation of outbreak control activities. This classification was used to calculate timeliness metrics for each stage, defined as the median time in days between two consecutive milestones, as well as responsible actors for each milestone. We also reviewed national guidelines (Ministerio de Salud (MINSA), 2017) to determine the metrics used for each process and aligned them with the Outbreak Milestones Framework.

**Table 1:**
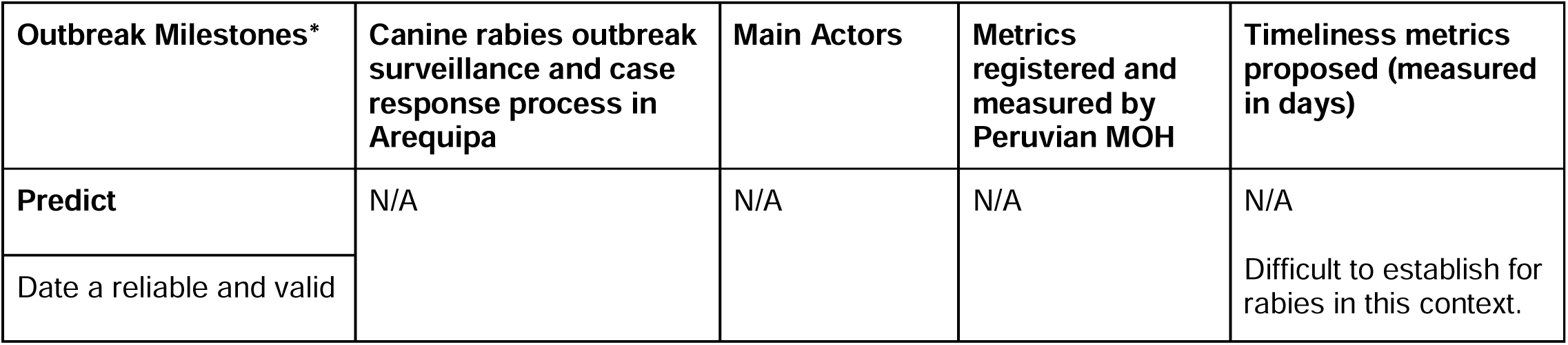
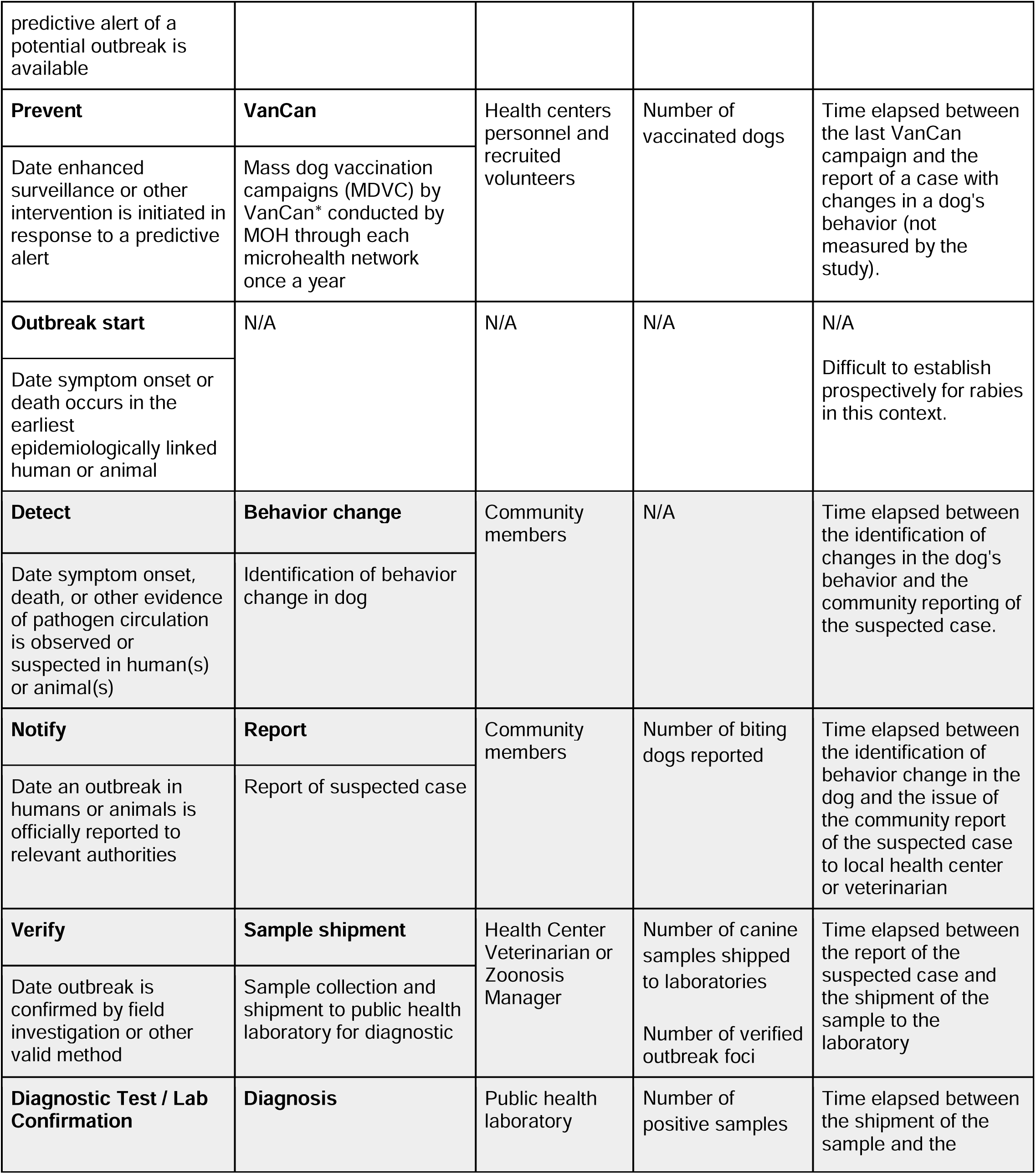
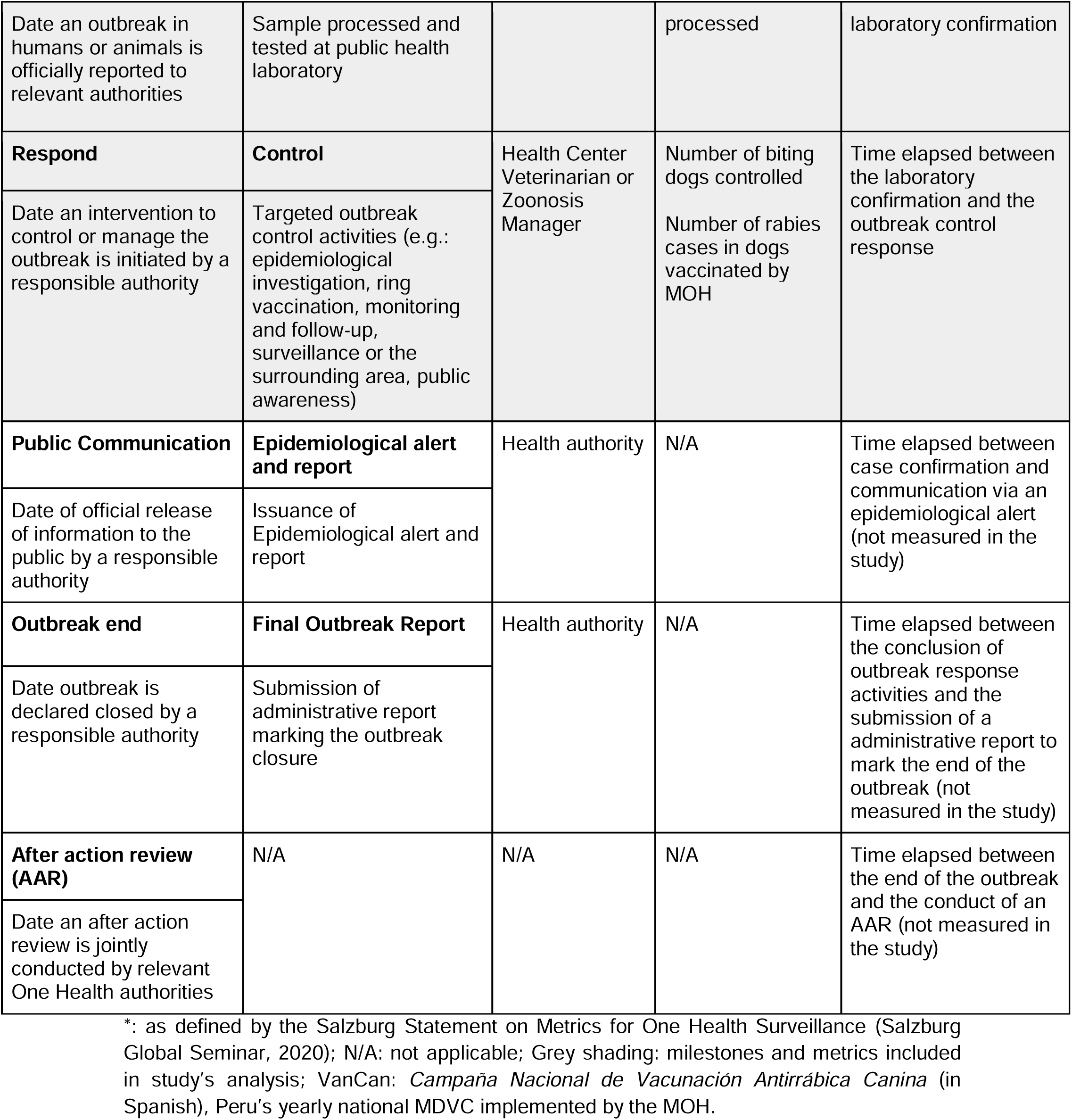
Classification of Canine Rabies Response Processes, Key Actors, and Timeliness Metrics Based on the Outbreak Milestones Framework.

**Table 2:** Comparison of Process Delays by Year Using Dunn’s Test.

| Timeliness Metric | Years Compared | p-value |
| --- | --- | --- |
| Symptom Onset Report - Sample Shipment | 2017 - 2021 | 0.003 |
| Symptom Onset Report - Sample Shipment | 2019 - 2021 | 0.015 |
| Symptom Onset Report - Sample Shipment | 2017 - 2022 | 0.005 |
| Symptom Onset Report - Sample Shipment | 2019 - 2022 | 0.023 |
| Sample Shipment - Laboratory | 2015 - 2018 | 0.004 |
| Sample Shipment - Laboratory | 2015 - 2021 | 0.002 |
| Sample Shipment - Laboratory | 2015 - 2022 | 0.001 |
| Sample Shipment - Laboratory | 2015 - 2023 | 0.005 |
| Laboratory - Control | 2016 - 2018 | 0.045 |
| Laboratory - Control | 2016 - 2022 | 0.014 |
| Laboratory - Control | 2016 - 2023 | 0.046 |

### Data Management and Analysis

#### Calculation of Timeliness Metrics and Statistical analysis

Following the process classification, timeliness metrics were calculated by establishing the time in days elapsed between two consecutive processes or milestones. We calculated four metrics between five milestones in all cases for 1) the time between behavior change and reporting of symptoms, 2) the time from reporting to shipment of the sample, 3) the time from sample shipment to receiving a laboratory confirmation, and 4) the time between getting laboratory confirmation to the initiation of outbreak control activities.

All statistical analyses were conducted using R version 4.4.3. To describe delays for each timeliness metric, numerical variables (delays by process) were summarized using the mean and median as measures of central tendency, and the standard deviation and interquartile range as measures of dispersion. Density plots and histograms were used to visualize differences by year and micro-network. The Shapiro-Wilk test indicated that the data followed a non-parametric distribution; therefore, the Kruskal-Wallis test was applied to compare delays across years and micro-health networks of the Arequipa Health Network. Additionally, the Spearman correlation coefficient was used to analyze the relationship between different types of delays.

#### Qualitative analysis of field report entries

We analyzed the digitalized narrative entries from all outbreak case reports in detail to identify systematically the main factors contributing to response delays. Based on the initial reading of reports and the outbreak response activities, we developed a codebook using a hybrid of both deductive codes (steps of the process) and inductive codes (emerging themes). A team of three researchers independently coded all reports using Dedoose software version 9.0.107 (*Dedoose Version 9.0.107, Web Application for Managing, Analyzing, and Presenting Qualitative and Mixed Method Research Data*, 2023) to ensure consistency and reliability in the analysis.

## RESULTS

We analyzed the reports of positive canine rabies cases in Arequipa (2015–2024) to evaluate delays across key outbreak surveillance and response milestones, aligning activities to the Outbreak Milestones framework (Table 1). We matched eight out of the eleven milestones, and proposed timeliness metrics to canine rabies control. Due to data limitations, we calculated only five timeliness metrics based on intervals between five milestones. Although MOH guidelines (MINSA, 2017) define metrics for five milestones, these focus primarily on process outputs and reporting volume rather than timeliness. Of the 392 confirmed cases reported during the study period, we excluded 71 cases (18.2%) due to incomplete or missing data. The final analysis was conducted with 319 confirmed cases.

The time interval between behavior change and symptom onset reporting shows a mean delay of 2.52 days (SD = 2.28) and a median of 2 days (IQR = [1 – 3]), with a relatively dispersed distribution (Figure 4). In contrast, the interval between laboratory diagnosis to initiation of outbreak control activities showed a shorter and more concentrated and consistent delay, with a mean of 0.76 days (SD = 0.92) and a median of 1 day (IQR = [0 – 1]) (Figure 4).

**Figure 4:**
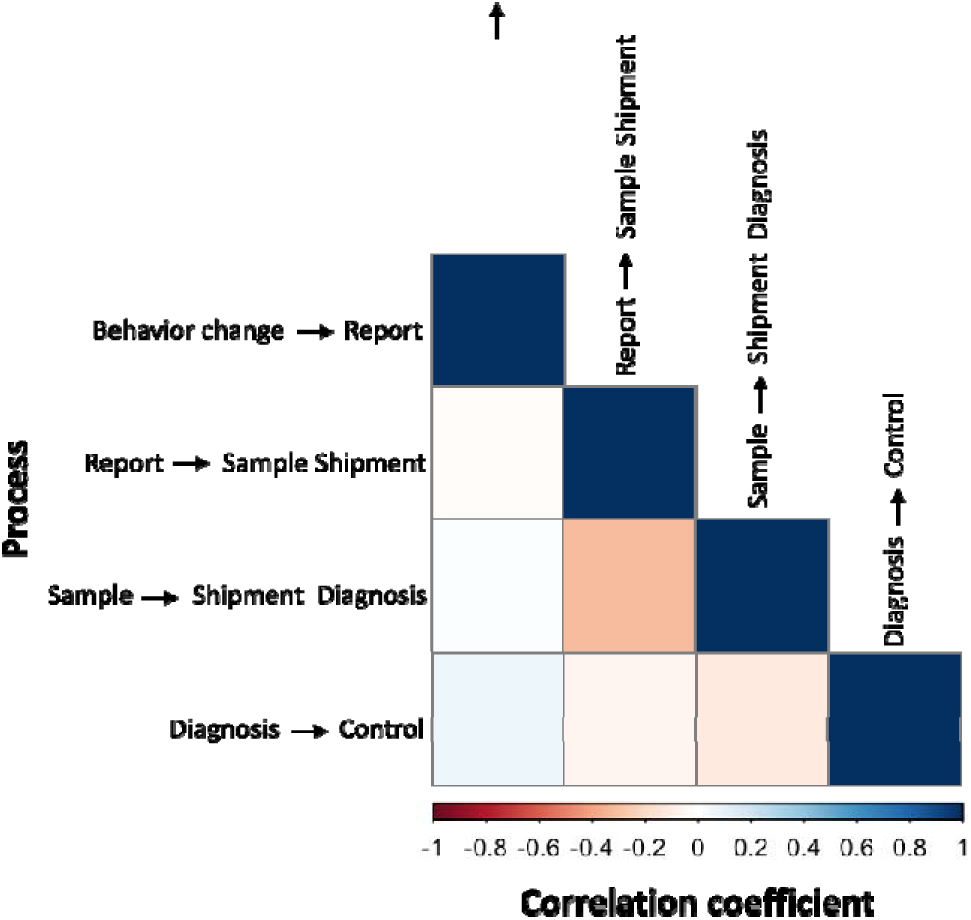
Correlation Coefficient between each Process in the Canine Rabies Surveillance System in Arequipa (2015–2024).

The interval between suspected case reporting and sample shipment showed a mean delay of 1.28 days (SD = 1.30) and a median of 1 day (IQR = [0 – 2]). Meanwhile, the interval between sample shipment and laboratory diagnosis has a mean of 1.09 days (SD = 1.38) and a median of 1 day (IQR = [0 - 1]). Variability in delays decreases as the average delay shortens, reflecting greater temporal consistency in these shorter delays. These findings suggest an inverse correlation between delay duration and operational consistency across time intervals, suggesting that improvements in process management and optimization could reduce both the length and variability of delays.

Correlation analysis between the different process time intervals (Figure 4) revealed very weak and non-statistical significance (p ≥ 0.05) association. However, a weak negative correlation was observed in the time interval between reporting and sample collection and the time interval between sample collection and diagnosis (r = -0.31, p < 0.05), suggesting an inverse association between these time intervals.

The delays in each time interval were compared by year from 2015 to 2024 (Figure 5). In the first time interval (between onset of symptoms and reporting the case to authorities), the distribution of mean delays varied between 2.00 days and 3.16 days. The years with the highest reported delays were 2017 (3.10 days) and 2023 (3.16 days), while the lowest delays were observed in 2015 (2.00 days) and 2021 (2.29 days). The dispersion of data in this time interval was the highest compared to the other time intervals.

**Figure 5.**
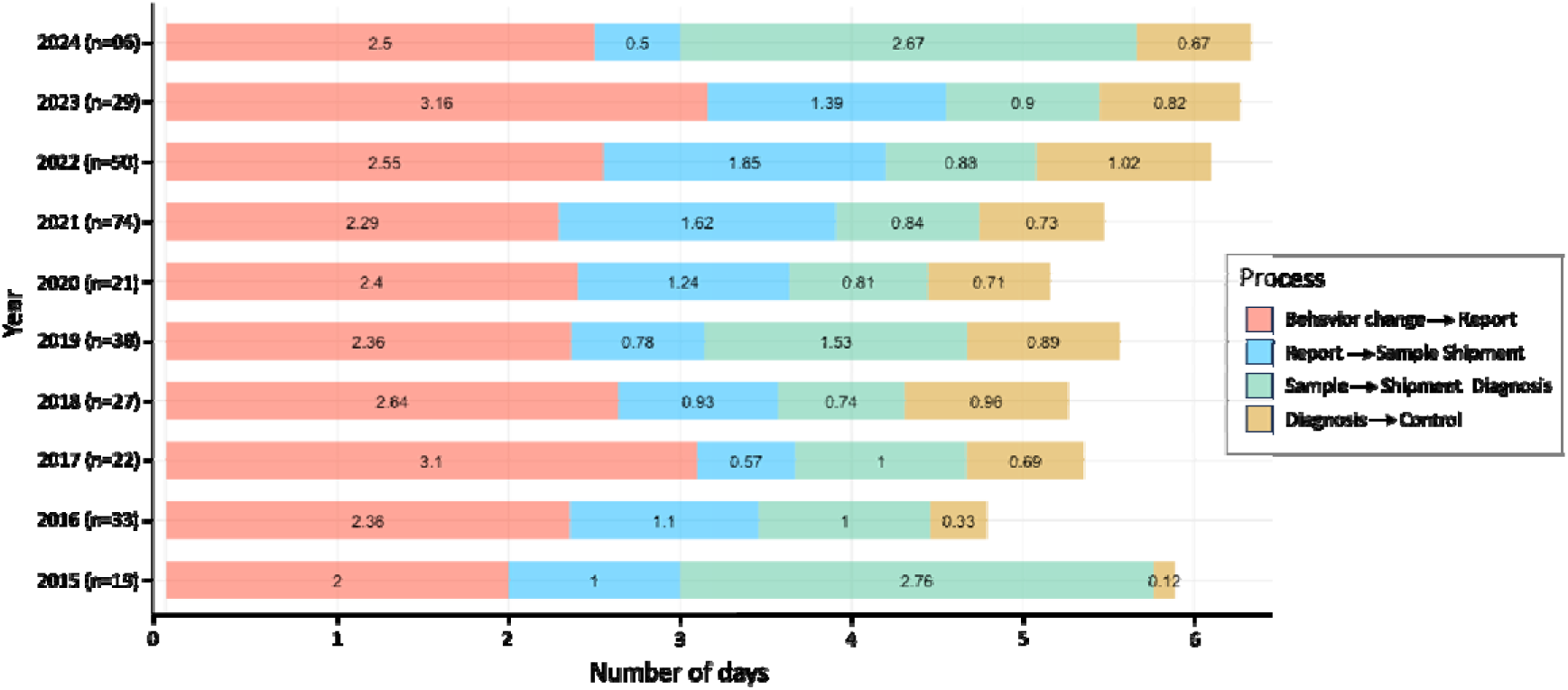
Average time spent (days) between each step of the rabies surveillance system per year, between 2015 and 2024.

The time interval between the reporting to the sample shipment shows distributions that vary between 0.5 days and 1.65 days. The years with the highest observed delays were 2021 (1.62 days) and 2022 (1.65 days), while the lowest delays were observed in 2017 (0.57 days) and 2024 (0.50 days). This time interval shows variations in the delay trends, with values tending to decrease from 2015 to 2019 and then increase from 2020 to 2022.

The interval between shipping the sample to laboratory diagnosis shows distributions that vary between 0.47 days and 2.76 days. The longest delays were observed in 2015 (2.76 days) and 2024 (2.67 days), while the shortest delays occurred in 2018 (0.74 days), 2020 (0.81 days), and 2021 (0.84 days). This time interval shows greater variability, with substantial increases in delays in some years and reductions in others.

Similarly, the time interval from the time of obtaining a diagnosis to outbreak control activity shows distributions that vary between 0.12 days and 1.02 days. The highest delays were seen in 2018 (0.96 days) and 2022 (1.02 days), while the lowest delays were observed in 2015 (0.12 days) and 2016 (0.33 days). This time interval continues to show a more consistent delay distribution compared to the other time intervals.

Regarding total delays by year (the sum of all four time intervals), the greatest total delay occurred in 2024 (6.34 days), while the lowest total delay was recorded in 2016 (4.79 days). The trend shows a progressive increase in total delays from 2016 to 2024, with a notable rise between 2022 and 2024.

Significant differences in delays across years were observed for time intervals from symptom onset reporting all the way to initiation of outbreak control measures. Kruskal-Wallis test revealed significant differences for the time between symptom onset reporting to sample shipment (p < 0.05), sample shipment to laboratory diagnosis (p < 0.05), and diagnostic confirmation to outbreak control activity (p = 0.006). These findings indicate that, in at least one year per process, the delay times differed significantly by year.

Post-hoc analysis using the Dunn test identified specific years with significant differences in delays for each process time interval. In the second process, significant differences were found between 2017 and 2021 (p = 0.003), 2019 and 2021 (p = 0.015), 2017 and 2022 (p = 0.005), and 2019 and 2022 (p = 0.023). For the third process, significant differences were observed between 2015 and 2018 (p = 0.004), 2015 and 2021 (p = 0.002), 2015 and 2022 (p = 0.001), and 2015 and 2023 (p = 0.005). In the fourth process, significant differences were found between 2016 and 2018 (p = 0.045), 2016 and 2022 (p = 0.014), and 2016 and 2023 (p = 0.046).

Delays for each metric were also measured across the 21 micro-health networks. The analysis excluded three micro-health networks as they only reported one case each during the whole study period. Initial reporting of symptom onset (i.e., behavior change in dog) ranged from 1.20 to 4.00 days, showing the highest variation of delays across micro-health networks or “microredes”. Symptom onset reporting to sample shipment revealed a heterogeneous trend, with some microredes presenting short delays of 0.50 days and others up to 2.2 days. This trend was also apparent in the interval between sample shipment to laboratory diagnosis, with delays ranging from 0.25 to 2.89 days. The final stage of diagnostic confirmation to initiation of outbreak control showed consistent distributions among microredes ranging from 0.25 to 2 days. Total delays for the whole process across microredes ranged from short spans of 3.43 days up to 7.85 days. Kruskall-Wallis test showed no significant differences in delays across microredes for any of the processes between outbreak milestones, suggesting that the observed variations are either not substantial enough or are masked by internal variability within microredes.

### Qualitative analysis of outbreak reports

Our qualitative analysis of the case report narratives about all targeted outbreak response activities was conducted to identify key factors contributing to delays. The reasons for each delay which were identified for each process (symptom onset in dog, report, diagnosis, and control activity) were coded and extracted from field reports, and are summarized below (Table 3). Notable excerpts illustrating some of the reasons are detailed below.

**Table 3:**
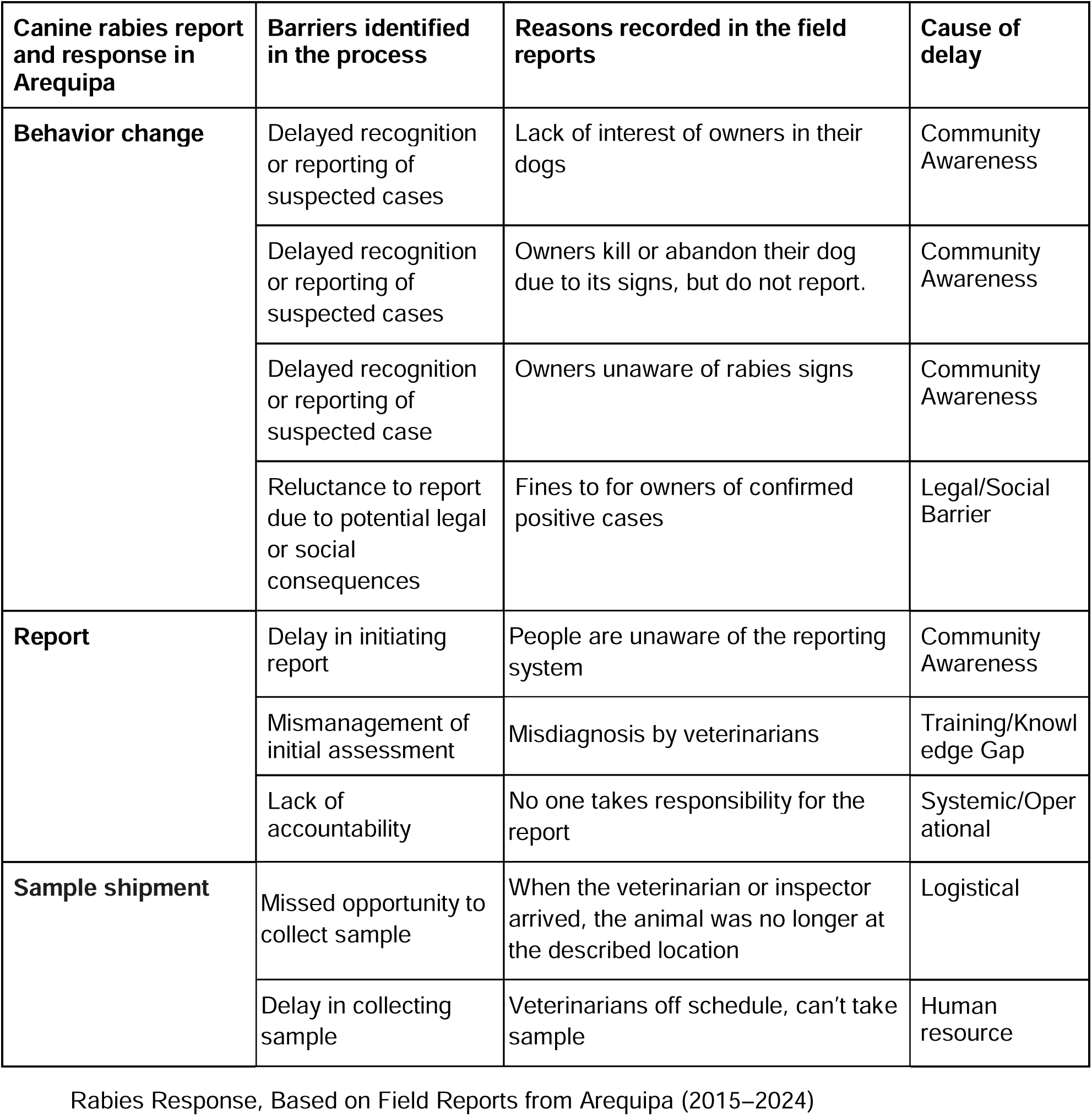
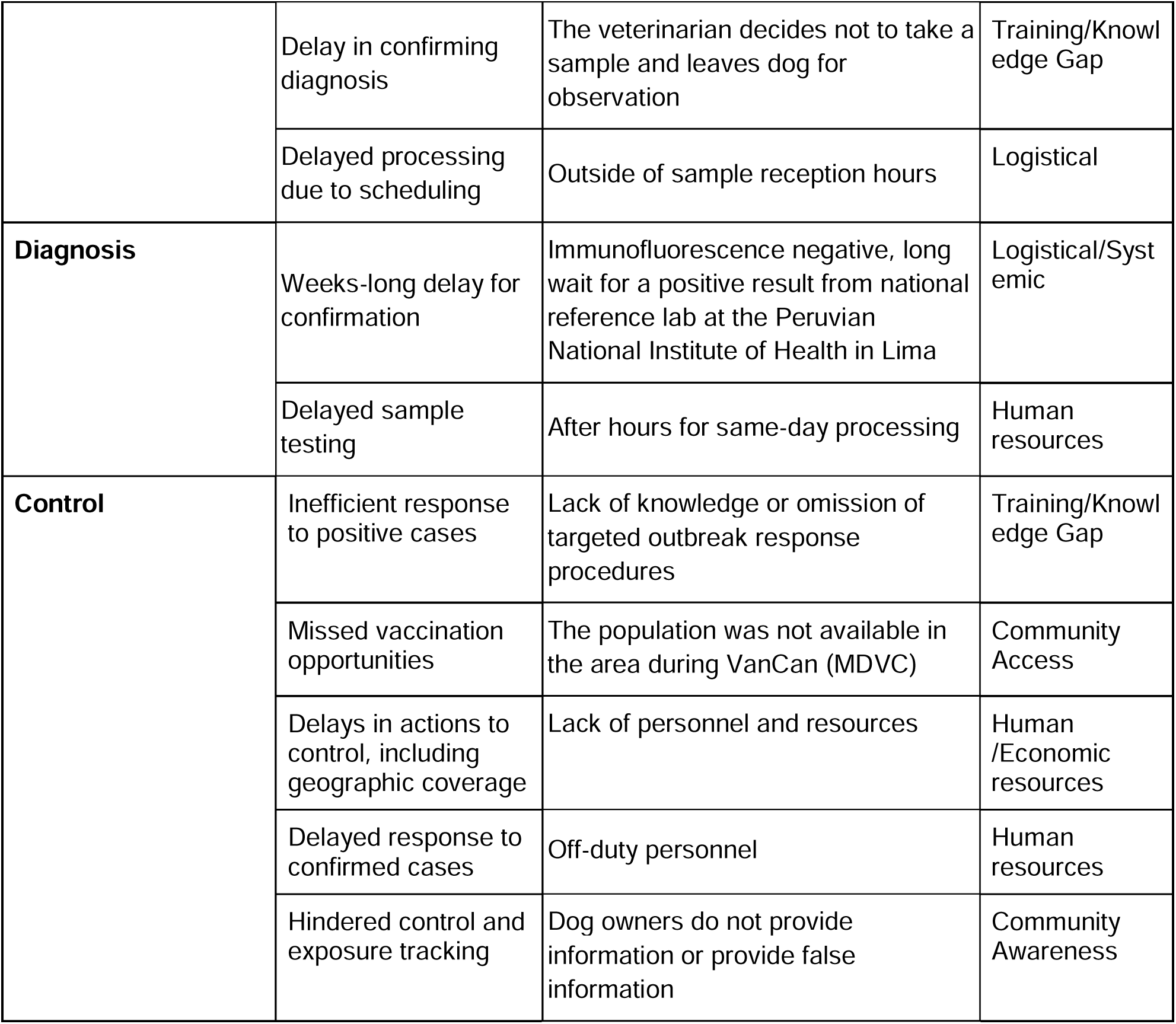
Identified Barriers and Causes of Delay Across Outbreak Milestones in Canine.

| Canine rabies report and response in Arequipa | Barriers identified in the process | Reasons recorded in the field reports | Cause of delay |
| --- | --- | --- | --- |
| **Behavior change** | Delayed recognition or reporting of suspected cases | Lack of interest of owners in their dogs | Community Awareness |
|  | Delayed recognition or reporting of suspected cases | Owners kill or abandon their dog due to its signs, but do not report. | Community Awareness |
|  | Delayed recognition or reporting of suspected case | Owners unaware of rabies signs | Community Awareness |
|  | Reluctance to report due to potential legal or social consequences | Fines to for owners of confirmed positive cases | Legal/Social Barrier |
| **Report** | Delay in initiating report | People are unaware of the reporting system | Community Awareness |
|  | Mismanagement of initial assessment | Misdiagnosis by veterinarians | Training/Knowledge Gap |
|  | Lack of accountability | No one takes responsibility for the report | Systemic/Operational |
| **Sample shipment** | Missed opportunity to collect sample | When the veterinarian or inspector arrived, the animal was no longer at the described location | Logistical |
|  | Delay in collecting sample | Veterinarians off schedule, can't take sample | Human resource |
| Rabies Response, Based on Field Reports from Arequipa (2015–2024) |  |  |  |
|  | Delay in confirming diagnosis | The veterinarian decides not to take a sample and leaves dog for observation | Training/Knowledge Gap |
|  | Delayed processing due to scheduling | Outside of sample reception hours | Logistical |
| **Diagnosis** | Weeks-long delay for confirmation | Immunofluorescence negative, long wait for a positive result from national reference lab at the Peruvian National Institute of Health in Lima | Logistical/Systemic |
|  | Delayed sample testing | After hours for same-day processing | Human resources |
| **Control** | Inefficient response to positive cases | Lack of knowledge or omission of targeted outbreak response procedures | Training/Knowledge Gap |
|  | Missed vaccination opportunities | The population was not available in the area during VanCan (MDVC) | Community Access |
|  | Delays in actions to control, including geographic coverage | Lack of personnel and resources | Human/Economic resources |
|  | Delayed response to confirmed cases | Off-duty personnel | Human resources |
|  | Hindered control and exposure tracking | Dog owners do not provide information or provide false information | Community Awareness |

#### Behavior Change

A recurring theme was that many owners either failed to recognize the behavior change or did not prioritize the health of their pets, sometimes lacking indifference toward their animals, which led to delays in identifying and addressing potential rabies cases. In one case report, it was noted: “*The owner indicated that she did not give importance to the dog’s behavior [change]*”. Even as signs of rabies became clearer, , some owners rather than consulting authorities or reporting the issue, some owners opted to either kill their pets or abandon them which not only exacerbated the problem but also increased the risk of exposure to other people and animals: “*Then, the dog was thrown into the street, where a day later it was seen 8 blocks away in the arms of a girl who picked it up to adopt it*.” Sometimes dog owners did express concern about their doǵs condition, but failed to recognize the behavior change as a sign of rabies, mistaking it for other illnesses or conditions. One field report described:

“On Monday [*date*] he began to show signs of paralysis and convulsions, the owners thought he was poisoned and treated him at home with oil. Since he had no signs of improvement, they strangled him.

##### Report

A recurring barrier identified to making a report was the lack of community awareness about the appropriate reporting system for suspected rabies cases. In some of the cases, community members observed dogs displaying unusual behavior but were uncertain about whom to notify. As the report recorded:

> *“(…) they report that the black and white dog came in on Saturday morning, [date] behaving strangely: He was disoriented, was uncoordinated when walking, attacked other dogs, but it is not clear if he bit the dogs in the [car mechanic] shop. People did not know who to report the case to.”*

In some cases, owners tried to be proactive about their concerns, but their lack of knowledge about proper procedures led to missteps, some that could even be dangerous. One owner euthanized their dog and collected a blood sample for analysis:

> *“The owner decided to euthanize the dog and collected a blood sample, then went to the [locality name] hospital for testing. There, they explained to him that this was not the procedure to follow and told him that the brain would have to be extracted at the local health center for testing.”*

Some owners reached out to their local health centers or authorities, but encountered staff and authorities that were also unsure about next steps or who did not want to take responsibility for next actions, resulting in a cycle of referrals to different institutions without resolution (which can also explain why some owners may not have bothered to report unusual dog behaviors to start with). One reported case captured the frustration of a dog owner:

> *“The next day at dawn the dog began to howl in a strange manner, the owner went to the health center in [district name] and there they told him that they did not provide that type of service and directed him to the police station. The owner went to the police station and they referred him to the security guard, and finally the security guard referred him back to the health center. The owner felt that they were making him go from one place to another without any solution [being ‘kicked around’] and he decided to euthanize the dog and bury it.”*

##### Sample shipment

Several barriers were found in the process of taking and shipping samples to the laboratory for testing. One common issue was the inability to locate the animal at the reported site. Sometimes the dog had left or escaped, while in others, the dog’s buried body was no longer present where indicated, often due to scavenging by other animals. Another significant obstacle was the lack of necessary supplies for euthanizing dogs, which hindered timely sample collection. Delays were also caused by decisions to leave animals under observation at their owner’s homes instead of taking immediate samples. One report described:

> *“On Wednesday the owner kicked the dog in the afternoon, he [the dog] was sad, ate little, drank water. On Thursday, the first observation was made at his home, he was apparently healthy and recommendations were given in case the dog disappears or dies. On Monday at 3:00 am the dog dies and [woman] informs [health inspector name] about the dog’s death and at 9:00 am the encephalic sample is taken.”*

Scheduling challenges further contributed to delays. Veterinarians were often called after hours, postponing sample collection and shipment to the following day when they were back on duty. In some cases, samples were taken outside the laboratory’s regular reception hours, causing additional delays in submission:

> *“The veterinarian [name] commented that he received the call from veterinarian [name] between 17:00 - 18:00 pm. Because it was late, the brain sample was taken the next day, Saturday [date]”*
>
> *“[Woman’s name] reported what had happened to [veterinarian’s name] who came to the home and took the sample at approximately 3:00 p.m. He didn’t send it to the lab because it was late. On Monday, [date], he [the veterinarian] took the sample [to the lab] at noon.”*

##### Diagnosis

Technical and procedural barriers delayed the process of diagnosing suspected rabies cases. Negative results of local immunofluorescence tests require confirmation at the national reference laboratory in Lima, which may introduce weeks of delay in responding to cases that were classified as negative locally. One field report described such a case:

> *“On November 26, the sample submitted was negative, and it was sent to the INS [National Institute of Health of Peru] for verification. On December 12, they confirmed it was a positive case, and on December 13, the corresponding outbreak control was conducted.”*

##### Control

Rabies control efforts also revealed challenges in implementing targeted outbreak response procedures. One major issue was the lack of knowledge or the omission of standard control procedures among the personnel involved:

> *“When arriving to the location of the positive case, just then the transport (ambulance) went to the health post of [locality name] to request personnel (5) to support in the focal control. These [health] personnel were unaware of the rabies case and the [veterinarian name] had not coordinated with the health promotion area and [they] indicated that they would only support until 11:30 because they had other activities already scheduled. At 12:00 pm they had a monthly meeting.”*

Another recurring challenge was the absence of residents in the affected area, which limited the ability to vaccinate owned dogs against rabies. Reports noted that, despite efforts to locate residents, many were unavailable, leaving some dogs in the focal control area unvaccinated: *“During the outbreak control procedure, not many residents could be found in the area, so some owned dogs did not receive the rabies vaccine.”*

Resource and personnel shortages were also significant obstacles. In some cases, the radius of the targeted control area was reduced due to insufficient personnel, and vaccination efforts were delayed because transportation was not available at the designated times. Additionally, off-duty personnel often delayed targeted outbreak response activities:

> *“The outbreak control could not be carried out on the day the case was confirmed because the veterinarian in charge [name] was not on duty. On Friday, [date], we were informed of the positive case and were told to go and support first thing on Saturday morning. The next day, we arrived at the [locality name] Health Center at 7:50 a.m. to coordinate with [man’s name], but when we called him, he informed us that he was not scheduled to work that day and that, additionally, they did not have an ambulance or driver, and that the outbreak control would be carried out on Monday [date].”*

Finally, misinformation and lack of cooperation from dog owners impeded outbreak control efforts. Individuals who owned or were in contact with rabid dogs occasionally refused to disclose their addresses or allow health personnel to visit their homes. In certain instances, they provided false information, further complicating contact tracing and implementation of control measures:

> *“The young man who rescued the dog refused to provide his address or bring medical personnel to his home because he was a rescuer and had several animals at home, all of which had been exposed to the rabid animal. Currently, the citizen who took the rabid dog to the veterinarian has not been located (he did not provide complete information, and some of the information he did provide is inaccurate), nor has the address been found.”*

## DISCUSSION

Epidemiological surveillance systems are essential for the control and elimination of infectious and zoonotic diseases such as rabies, as they enable early detection and effective outbreak responses. However, the effectiveness of these systems depends on the timeliness and coordination of case detection, reporting and control measures. In Arequipa, Peru, dog-mediated rabies re-emerged and have persisted despite sustained control strategies. Using the Outbreak Milestones and Timeliness Metrics framework, this study identified and analyzed surveillance and control activities time intervals and the main contributing factors to delays in the detection, reporting and response to cases of canine rabies in Arequipa, Peru, between 2015 and 2024. Our findings reveal critical inefficiencies, particularly in community-level reporting and logistical coordination, hindering timely enaction of a control response. This study provides a blueprint basis for improving rabies control in Arequipa and may serve as a model for strengthening outbreak responses to other zoonotic diseases.

Mapping the rabies surveillance and response system allowed us to easily identify key processes and align them with their corresponding One Health Outbreak Milestones. This helped establish the specific timeliness metrics evaluated in this study. Although the framework recognizes that milestones may not occur in a strict sequence, we were able to classify the rabies processes in a consistent order. However, we acknowledge that certain steps, such as the sample shipment, diagnosis and initiation of control measures, can occur in parallel rather than sequentially, which may influence how delays are interpreted. We focused on systematically measuring the time intervals between processes related to the detection, reporting, confirmation and response of rabies cases. VanCan, the national MDVC, was identified as a prevention-related milestone. Future studies could explore the interval between the last MDVC and the emergence of new cases, which may provide insights into vaccination coverage and campaign timing.

Another relevant milestone that we did not explore is public communication. While we identified the release of epidemiological bulletins by the MOH as a proxy, we could not systematically measure the time between case confirmation and public reporting. Future work could focus on this milestone and how the case information reaches affected communities, or through which channels, and understand aspects of public trust and transparency. We did not further assess the Prediction milestone, as it remains particularly difficulty to establish for rabies in the studied context, due to multifactorial drivers of the disease ecology, as well as underreporting. We hope that predictive modeling could be achieved with future applications of genomic surveillance, spatial statistics and risk mapping that can be operationalized to support an early-warning system.

We were also not able to identify a public source for the official closure of outbreaks, preventing us from evaluating the End of Outbreak milestone. Exploring how closure declarations could be standardized, may provide clarity in outbreak management timelines and use of resources. The One Health Outbreak Milestones framework proposes a final post-response feedback milestone involving after-action reviews. These are not conducted at this level for dog-mediated rabies in Peru, but we believe that incorporating systematic feedback would strengthen coordination, accountability and learning. The timeliness metrics we propose could provide a basis for informing and structuring such future reviews.

The time between detection of behavior changes in dogs and reporting the onset of signs to health authorities represents the largest delay in the system. This indicates a bottleneck in the reporting process at the community level, which may contribute to persistence and spread of rabies as well as complicate control efforts, as evidenced. Lack of owner knowledge of rabies symptoms is a known cause for lags in case identification. In order to reduce these delays, increasing community education, awareness and engagement in health promotion campaigns, may improve the community’s ability to identify symptoms and their willingness to report suspected cases promptly.

The delays associated with the health system’s response to community reports reveal recurring logistical issues and a lack of available trained personnel. These limitations were noted consistently in both quantitative and qualitative analyses consistently detected, where administrative barriers and poor inter-institutional coordination were identified as critical factors affecting the speed of response. These types of delays in sample collection and transportation are well documented in zoonotic disease surveillance systems, and contribute to reduced efficiency in rabies response efforts. The variability in response times between health micro-health networks further highlight inequities in resource allocation, with periurban areas facing disproportionate delays due to lack of health services and limited access to veterinary care. Strengthening the health system’s capacity is critical to improving response times. Our findings heavily point towards implementing standardized response protocols and continuous personnel training to expedite sample collection and timely notification to laboratories. For instance, delays observed at this level in 2015 could be attributed to the early stages of implementing the post-reintroduction rabies surveillance system, similarly to Tanzania during the reactivation of anti-rabies programs, underscoring the need for ongoing investment in process improvement.

The delays between diagnosis and outbreak response focal align with findings in other LMICs, where poor inter-institutional coordination and insufficient logistical resources significantly delayed response times. Our qualitative analysis revealed that lack of standardized protocols and ineffective communication between health authorities and affected communities further contributed to these delays.

A marked variability in system response delays across years was observed, with periods of greater delay associated with years of public health budget cuts or disruptive events, such as the COVID-19 pandemic, particularly hindering MDVC in areas severely affected by rabies, due to the overload of the health system and the reassignment of resources for human emergency care. These variations support the case for sustained investment in prevention and preparedness to reduce system vulnerabilities and achieve consistent response times. At the micro-health network level, disparities were more pronounced, particularly between urban and periurban areas in the city of Arequipa. Urban micro-health networks, with greater access to resources, experienced fewer delays compared to their periurban counterparts, where issues like transportation barriers and staff turnover exacerbated the delays by almost doubling them. Discrepancies in the post hoc analysis suggest the gaps are not systematic, but localized challenges persist due to fragmentation and perpetuated inequalities, where focused training reduced these disparities, calling for differentiated policies with targeted interventions to address area-specific needs, emphasizing resource-limited districts and networks. Micro-health networks with proactive leadership and community coordination were more equipped to overcome these barriers, emphasizing the role of local governance in adapting to adverse conditions.

The variability found in delays across the system highlights the importance of an equitable approach in resource allocation, ongoing staff training, and the standardization of operational protocols across all micro-health networks. These findings underscore the need for continuous monitoring and adaptive that can respond to changing conditions and reduce disparities in response times. Adaptive, resilient and sustainable systems are key to the response of any public health emergency. Our results provide evidence for prioritization of interventions at certain levels. Implementing targeted strategies could help standardize response times and reduce disparities between health micro-networks. Ultimately, integration of approaches based on community education, building sustainable local capacities, and logistical optimization, will be key to enhancing early-warning efficiency of the surveillance system.

While our study reveals critical delays in the system that need to be addressed, it also highlights missed opportunities for assessing the performance and quality of the rabies public health system response. Peru’s MOH monitors output-based metrics, such as the number of samples collected or number of outbreaks reported, to evaluate some of the processes in the system, which may end up masking inefficiencies. Aligning these metrics with the different outbreak milestones, revealed gaps that limit the system’s ability to assess performance. Measuring beyond task completion, but response quality (accuracy of diagnosis, outbreak containment covered area) or timeliness, remain essential for a structured and clear understanding of the health of the surveillance system itself. Long-term improvement of outbreak preparedness and response for a robust, sustainable and resilient system may only be possible with feedback and accountability mechanisms in place.

This study is not without limitations. First, it is a secondary analysis using retrospective longitudinal data collected by various individuals over several years, introducing potential inconsistencies in record keeping and data quality. The lack of standardization in documentation across time and personnel may have influenced the accuracy of time interval measurements, particularly between the 2015-2018 period and the 2019-2024 period. Additionally, the exclusion of incomplete cases, though limited in number, could introduce a selection bias, and reduce the representativeness of the findings.

Future research should consider prospective data collection using standardize tools and mixed-method approaches. Community-participatory methods could help contextualize public reticence or delays in reporting. Operational research and human systems engineering models may assist in identifying bottlenecks and more granular barriers in workflows, and facilitate the design of sustainable solutions, while economic evaluations such as cost-benefit and cost-effectiveness analysis could support resource allocation decision-making. Evaluating the components and impact of health communication and education strategies may also be relevant, considering the canine rabies epidemic has been ongoing in Arequipa for a decade, as this may provide insights into how to better shape the strategy. Ultimately, mathematical modelling of outcomes considering delay variation could represent a valuable tool for more robust data-driven system change. Despite our results only being limited to Arequipa. Our findings are in tune with other studies in other diseases and areas of public health, highlighting the need for public health officials and institutions to focus on the systemic barriers the represent causes of delay like education, training, logistic, human resources, and economic resources.

## CONCLUSIONS

We identified significant delays in the canine rabies surveillance and response system in Arequipa. The analysis shows that the causes of delays are multifactorial and dynamic, with critical challenges in early detection by the community through focused strategies and in the operational response of the health system. These findings highlight the urgency of strengthening the articulation between the community and the health system, as well as optimizing resources to reduce the risk of secondary transmission and move towards the elimination of canine rabies. Our findings demonstrate the utility and importance of establishing and measuring timeliness metrics and the Outbreak Milestones framework for evaluating and optimizing public health responses. Further research, including prospective studies and community-based perspectives, will help refine these strategies and improve rabies control efforts in Arequipa and similar regions. Addressing these delays and their causes could help streamline surveillance and response actions. This study represents a real-world longitudinal case analysis of how the outbreak milestones and timeliness metrics can be identified, measured and dissected for surveillance of a zoonotic disease in an urban environment.

## SUPPORTING INFORMATION

COREQ checklist for qualitative data

## DATA AVAILABILITY

Databases and code used (github repo)

## ACKNOWLEDGMENTS

We would like to thank field personnel from the Zoonotic Disease Research Laboratory (ZDRL) for carrying focus control activity reports, and Gerencia Regional de Salud de Arequipa (GERESA) for facilitating our long-standing collaboration in the study of rabies in the city. We also want to thank and recognize the work of all of those involved in the rabies surveillance system in Arequipa, from the community, to our colleagues in the health centers, diagnostic laboratories and on the field.

